## Supplementary Material for "Social connections are differentially related to subjective age and physiological age acceleration amongst older adults"

[Supplementary Figure 8: relationship between social deficits and subjective and physiological age acceleration stratified by age (top panel, age<65; bottom panel age>=65) (For ages <65, subjective age n=3,261, physiological age n=2,118; for ages >=65, subjective age n=3,216, physiological age n=1,995)	12](#_Toc210054748)

### Appendix 1: Supplemental methods

#### Appendix 1a. Procedures for biomarker collection and inclusion criteria

Methods for blood-based biomarker processing are in line with standard procedures and described in detail elsewhere.1 Participants with CRP concentrations >20 mg/l were excluded as this can indicate acute infection.2 Reproduced from ELSA nurse visit documentation. Available in full at <https://www.elsa-project.ac.uk/study-documentation>.

**Blood pressure -** All respondents were eligible to have their blood pressure measured. Three measurements were taken of systolic and diastolic pressure as well as pulse rate on the respondent’s right arm while they were seated. The mean value of the valid measurements was included in the analyses. Readings were considered invalid if the participant had eaten, drank, smoked, or exercised in the last 30 minutes.

**Grip strength -** All respondents were eligible to have their grip strength measured. Grip strength was evaluated using a Smedley dynamometer. Three measures of grip strength in each hand were taken, with the maximum value in the dominant hand retained for analysis.

**Waist circumference -** All respondents were eligible to have their waist measurements taken, unless they were chair-bound or had a colostomy or ileostomy. Measurements were taken twice; however, if the second measurement differed from the first by 3cm or more, the nurse received an error message in the CAPI program and was prompted to either amend one of the previous responses if a mistake had been made entering a measurement, or to take a third measurement. If the nurse believed that the measurements they took were 0.5cm more or less than the true measurement because of problems encountered (e.g. clothing the respondent was wearing), this was considered unreliable.

**Blood samples -** All sample members who gave consent were eligible for a blood sample to be taken. The only exceptions to this were people with clotting or bleeding disorders, people with a history of fits or convulsions, or people who were on anticoagulant drugs. Respondents aged 80 or under were asked to fast before their nurse visit so a fasting blood sample could be taken. Respondents were not asked to fast if they had diabetes and were on treatment or if they were considered to be malnourished or otherwise unfit to fast (this information was obtained from the interviewer). Respondents who were asked to fast were given guidelines about when and what they could eat based on their appointment time. In the nurse visit, respondents were asked when they had last eaten and, if this was in the last 24 hours, what they had eaten. The CAPI program used their responses to work out if they had fasted adequately. A respondent was considered to have fasted and therefore be eligible for a fasting blood sample if they hadn't eaten or drunk anything (apart from water) on the day of their nurse visit OR they hadn't eaten or drunk anything (apart from water) in the past 5 hours and had only had a light meal (see appointment record card) or a piece of fruit or drink the last time they ate.

**Lung function -** Three measurements each were taken of forced vital capacity and forced expiratory volume using a NDD Easy On-PC spirometer, with the largest measure retained for analysis. All respondents were eligible to have their lung function measured, except for the following: (i) those who had had abdominal or chest surgery in the preceding 3 weeks (ii) those who had been admitted to hospital with a heart complaint in the preceding 6 weeks (iii) Those who had had eye surgery in the preceding 4 weeks.

#### Appendix 1b. Derivation of physiological age

Physiological age was derived based on the principal component analysis (PCA) method, which is among the most commonly used methods of biological age determination.^2–4^ The concept underlying this method is that the ageing of multiple organ systems can be represented as a linear combination of clinical markers that capture changes in organ function with age. By selecting clinical markers representing different organ systems that are associated with chronological age in a healthy subsample of the study population, PCA can be used to produce a score that represents the healthy ageing process of these organ systems. The weights derived from the healthy subpopulation are then used to produce physiological age in the wider study population.

Derivation of physiological age was performed using the original ELSA cohort respondents only without including refreshment cohorts (N=8,253). We first excluded 7,182 participants reporting diabetes, cancer, lung disease, cardiovascular conditions, stroke, arthritis, asthma, high blood pressure, high cholesterol, Parkinson’s disease, osteoporosis, or dementia, and 249 participants who were missing biomarker data, leaving 822 healthy participants in the derivation sample.

We standardised each biomarker by sex and examined correlations between the biomarker and chronological age at each wave, retaining those with a correlation coefficient of ≥0.10 at any wave. This led us to retain pulse, systolic blood pressure, fibrinogen, C-reactive protein, glycated haemoglobin, forced expiratory volume (FEV), forced vital capacity (FVC), and grip strength for further examination. We then performed PCA including chronological age and the retained biomarkers, predicted scores for the first component, and examined correlations between each biomarker and the predicted score. We retained biomarkers with at least moderate correlation (r≥0.3) with predicted score; we did not exclude any biomarkers at this stage. We performed PCA again with the remaining biomarkers but excluding chronological age. We used the first component to produce a physiological score for all participants, including those initially excluded due to chronic conditions. We converted this physiological score to physiological age using the following formula, which is the standard procedure for derivation of physiological age by PCA method:^2^

𝑃𝐴𝑖=(𝑃𝑆𝑖∗𝑠𝐶𝐴)+𝑥̄𝐶𝐴

Where 𝑃𝐴𝑖 denotes physiological age for individual 𝑖, 𝑃𝑆𝑖 denotes the physiological score for individual 𝑖, 𝑠𝐶𝐴 denotes the derivation sample standard deviation of chronological age and 𝑥̄𝐶𝐴 denotes the derivation sample mean chronological age. We applied the following correction to physiological age to reduce systematic error:^4,5^

𝑃𝐴𝑖=𝑃𝐴𝑖+[(𝐶𝐴𝑖−𝑥̄𝐶𝐴)×(1−𝑏)]

Where 𝐶𝐴𝑖 denotes chronological age for individual 𝑖, and 𝑏 denotes the simple linear regression coefficient of 𝑃𝐴 on 𝐶𝐴 in the derivation sample.

#### Appendix 1c. Outcome ascertainment and Cox proportional hazards models for internal validation

Participants were asked to report functional limitations lasting at least three months. ADL included walking across a room, dressing, bathing, eating, getting in/out of bed, and toileting. IADL included using a map, using the telephone, managing money, taking medications, grocery shopping, preparing a hot meal, and doing work around the house/garden. Mobility activities included walking 100 yards, sitting for two hours, getting up from a chair, climbing one flight of stairs, stooping/kneeling/crouching, lifting/carrying 10 lbs, picking up a coin, reaching/extending the arms, or pushing/pulling a large object. For a given wave, a participant was considered to have ADL limitations if they reported at least “some difficulty” with one or more ADL, IADL limitations if they reported “some difficulty” with one or more IADL, and mobility limitations if they reported “some difficulty” with one or more mobility activities.

To assess memory, participants were administered Consortium to Establish a Registry for Alzheimer’s Disease immediate and delayed recall tasks,^7^ in which participants are read a 10-word list and asked to recall it immediately and after a delay. Immediate and delayed recall scores are summed. For a given wave, participants were considered to show evidence of memory impairment if their memory score was >1.5 standard deviations below the mean for their five-year age group. This cut-off is widely used to identify cognitive impairment when clinical evaluation is not possible.^8^

At each interview participants were also asked to report whether they’d received a diagnosis of diabetes, lung disease, cardiovascular disease, stroke, high cholesterol, high blood pressure, cancer, arthritis, osteoporosis, dementia, or Parkinson’s disease.

Data from waves 2-10 were used to validate physiological ageing. Physiological age derived from the first biomarker measurement for each participant was used to produce ageing acceleration (physiological age – chronological age). We then used Cox proportional hazards models to examine associations between ageing acceleration and incident ageing-related health outcomes. These models had a time scale in years since measurement of ageing acceleration. The maximum follow-up period comprised nine waves or 19 years of follow-up (2004/05-2021/23). Separate models were fitted for each outcome. Multiple-event-per-subject models were used for functional limitations and memory impairment; single-event-per-subject models were used for chronic conditions. Data were censored at the interview of record of the outcome (single-event-per-subject only) or the end of follow-up (December 2021/23). Models were adjusted for chronological age and sex with the proportional hazards assumption evaluated using Schoenfeld residuals.

### Appendix 2: Supplemental tables

#### Supplementary Table 1. Components and indicators of social connections

| Social connections Component | Indicator | Questions |
| --- | --- | --- |
| Structural | Marital status | Current legal marital status |
|  | Intimate network size | How many of your children would you say you have a close relationship with? |
|  |  | How many of these family members would you say you have a close relationship with? |
|  |  | How many of your friends would you say you have a close relationship with? |
|  | Social integration | Are you a member of any of these organisations, clubs or societies? (political parties, trade unions, environmental groups, tenants/residents associations, Neighbourhood Watch, church or religious groups, charitable associations, evening classes, social clubs, sports clubs, exercise classes or other clubs/societies) |
|  |  | In the last 12 months, have you given any unpaid help to any groups, clubs or organisations? (raising money/sponsored events, committee membership/group leadership, organizing/running activity or event, visiting people, befriending/mentoring, education/teaching/coaching, providing information/counselling, secretarial/admin/clerical work, providing transport/driving, representing, campaigning, practical help like shopping, other help |
|  |  | In the last 12 months, how often have you been to the cinema, an art gallery, museum or exhibition, or the theatre, concert or the opera |
|  | Living alone | Number of people in the household |
|  | Social isolation  [children/relatives/friends] | How often do you meet up with them? |
|  |  | How often do you speak on the phone with them? |
| Functional | Loneliness | How often do you feel you lack companionship? |
|  |  | How often do you feel left out? |
|  |  | How often do you feel isolated from others? |
|  | Social support  [partner/children/relatives/friends] | How much do they let you down when you are counting on them? [reverse coded] |
|  |  | How much can you rely on them if you have a serious problem? |
|  |  | How much can you open up to them if you need to talk about your worries? |
| Quality | Negative aspects/strains  [partner/children/relatives/friends] | How much do they criticise you? |
|  |  | How much do they really understand the way you feel about things? [reverse coded] |
|  |  | How much do they get on your nerves? |

#### Supplementary Table 2. Associations between ageing acceleration and ageing-related outcomes

|  | **HR for 5-year change in**  **ageing acceleration (95% CI)** |  | *P-value* |
| --- | --- | --- | --- |
| *Functional limitations* |  |  |  |
| ADL | 1.15 (1.14-1.17) |  | <0.0001 |
| IADL | 1.16 (1.15-1.17) |  | <0.0001 |
| Mobility activities | 1.09 (1.09-1.10) |  | <0.0001 |
| Memory impairment | 1.16 (1.14-1.17) |  | <0.0001 |
| *Chronic conditions* |  |  |  |
| Diabetes | 1.34 (1.30-1.38) |  | <0.0001 |
| Lung disease | 1.23 (1.19-1.28) |  | <0.0001 |
| Cardiovascular disease | 1.09 (1.06-1.11) |  | <0.0001 |
| Stroke | 1.16 (1.12-1.21) |  | <0.0001 |
| High cholesterol | 1.08 (1.06-1.09) |  | <0.0001 |
| High blood pressure | 1.26 (1.23-1.28) |  | <0.0001 |
| Cancer | 1.03 (1.00-1.05) |  | 0.065 |
| Arthritis | 1.06 (1.03-1.08) |  | <0.0001 |
| Osteoporosis | 1.04 (1.01-1.07) |  | 0.018 |
| Dementia | 1.09 (1.05-1.14) |  | <0.0001 |
| Parkinson’s disease | 0.98 (0.90-1.07) |  | 0.63 |

Ageing acceleration corresponds to physiological age – chronological age. Based on Cox proportional hazards models adjusted for sex and chronological age. Outcomes ascertained from waves 2-10 of ELSA (2004/05-2021/23).
Abbreviations: HR, hazard ratio; CI, confidence interval; ADL, activities of daily living; IADL, instrumental activities of daily living.

#### Supplementary Table 3. Results of restricted cubic spline tests of nonlinearity for age-biomarker associations.

| **Biomarker** |  | **P-value*** |
| --- | --- | --- |
| Pulse pressure |  | 0.032 |
| Systolic blood pressure |  | 0.71 |
| Fibrinogen |  | 0.95 |
| C-reactive protein |  | 0.056 |
| Glycated hemoglobin |  | 0.16 |
| FEV |  | 0.42 |
| FVC |  | <0.0001 |
| Grip strength |  | <0.0001 |

Models are linear mixed models with a random intercept and slope at the individual level, fitted to biomarker data from waves 2, 4, and 6. P-values are from likelihood ratio tests comparing models with linear versus spline terms for age. Although several tests were statistically significant, spline plots (Supplementary Figure 1) indicated monotonic, approximately linear associations across the analytic age range.

#### Supplementary Table 4: Average treatment effects and 95% confidence intervals for main analyses

|  | Subjective age acceleration  (ATE + 95% CI; years) | Physiological age acceleration  (ATE + 95% CI; years) |
| --- | --- | --- |
| Living alone | -1.41 [-2.17,-0.66] *** | 2.58 [1.39, 3.77] *** |
| Small social network | -0.04 [-0.70, 0.62] | 0.90 [0.04, 1.77] * |
| Low social integration | 0.62 [-0.05, 1.29] | 1.88 [0.75, 3.01] *** |
| High social isolation | -0.94 [-1.69,-0.20] * | 0.82 [-0.23, 1.86] |
| Low social support | -0.46 [-1.12, 0.20] | 1.86 [0.91, 2.80] *** |
| High loneliness | 0.22 [-0.57, 1.01] | -0.54 [-1.77, 0.69] |
| High social strain | -0.10 [-0.75, 0.55] | -1.16 [-2.14, -0.17] * |

ATE = average treatment effect, CI = confidence interval. * = p<.05, ** p<.=01, *** p<.001

### Appendix 3: Supplemental figures

#### Supplementary Figure 1. Restricted cubic spline plots of age–biomarker associations (ages 50-90).

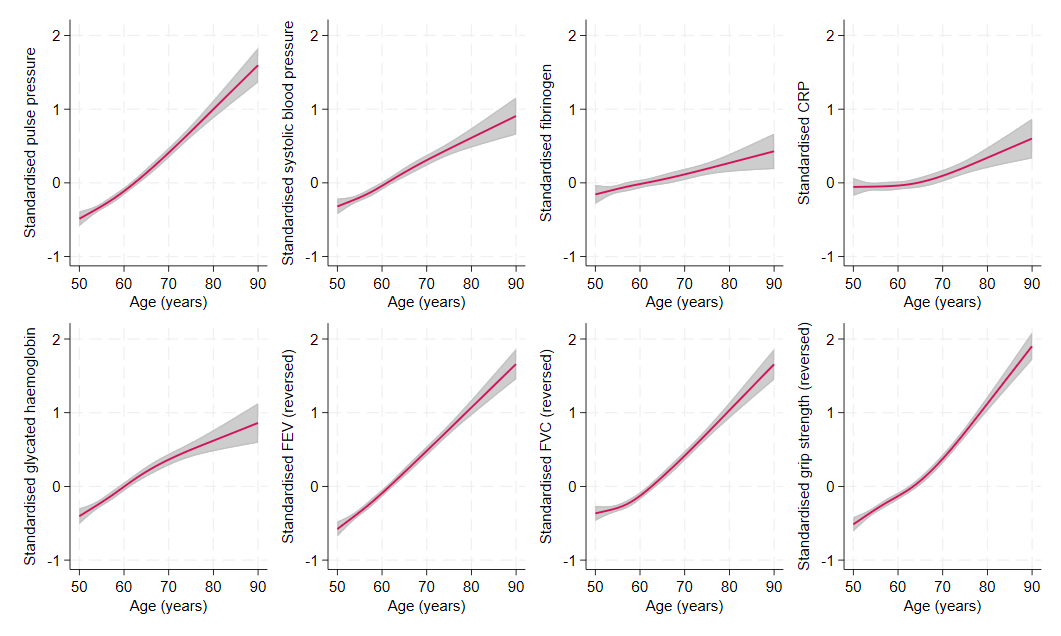

Plots based on linear mixed models with a random intercept and random slope for age at the individual level, fitted to biomarker data from waves 2, 4, and 6. Age was modelled using restricted cubic splines with four knots (at the 5th, 35th, 65th, and 95th percentiles of the analysis sample). Shaded areas represent 95% confidence intervals. Although likelihood ratio tests (Supplementary Table 3) indicated statistically significant departures from linearity for several biomarkers, the spline plots show monotonic, approximately linear associations across the analytic age range.

#### Supplementary Figure 2. Restricted cubic spline plot of the association between chronological age and the PCA-derived physiological age index (ages 50-90).

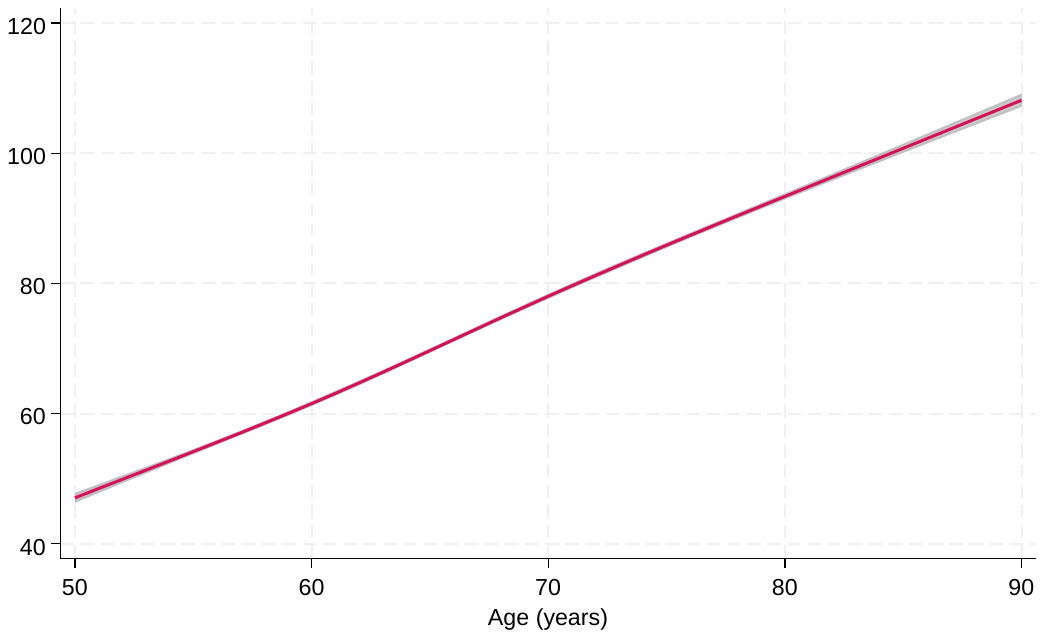

Plot based on linear mixed model with a random intercept and random slope for chronological age at the individual level, fitted to biomarker data from waves 2, 4, and 6. Age was modelled using restricted cubic splines with four knots (at the 5th, 35th, 65th, and 95th percentiles of the analysis sample). Shaded areas represent 95% confidence intervals.

#### Supplementary Figure 3: correlation matrix between all variables in the study

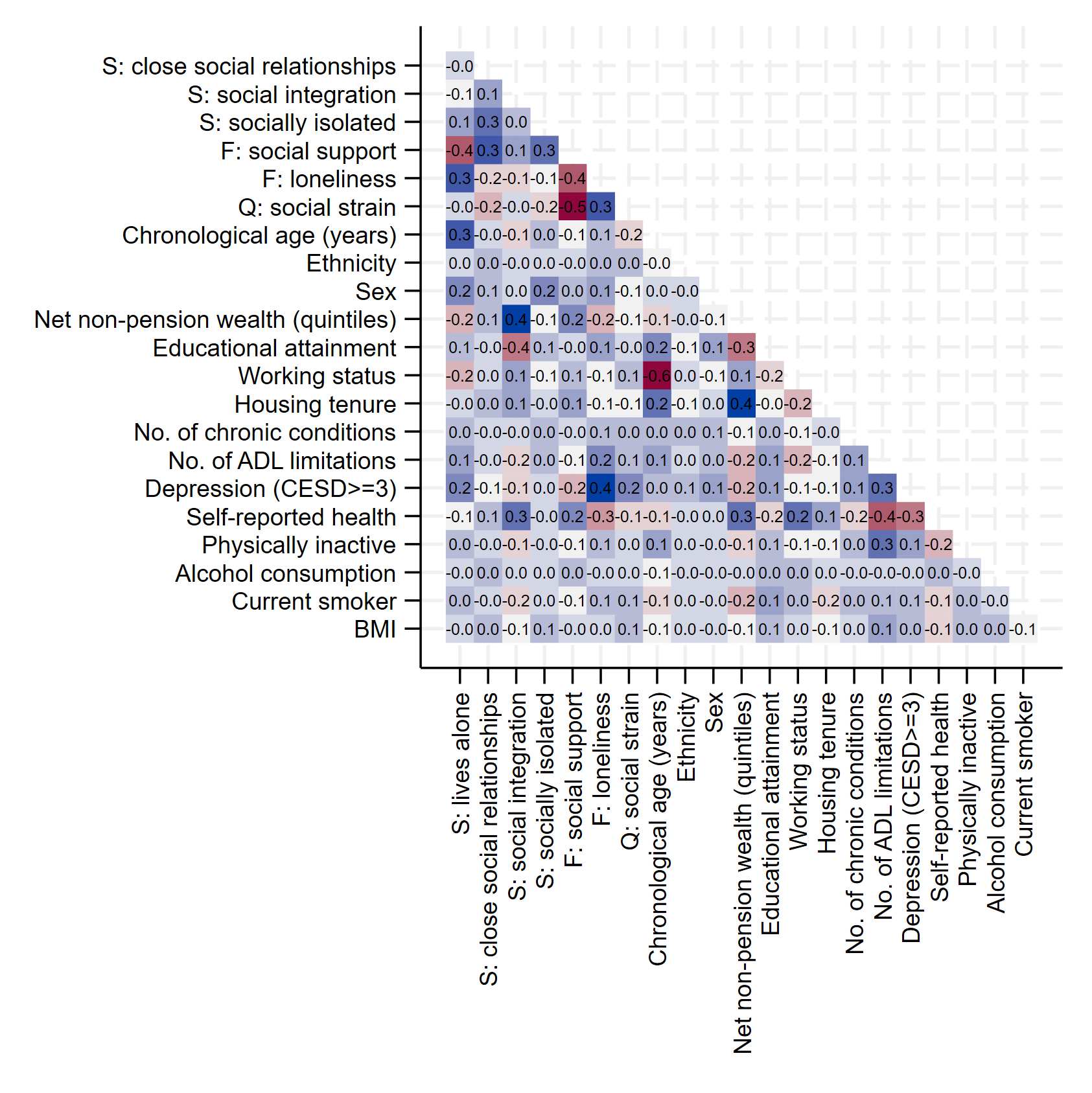

#### Supplementary Figure 4: bivariate histograms using hexagon heat plots to show the relative frequency of associations beween subjective and physiological age and chronological age

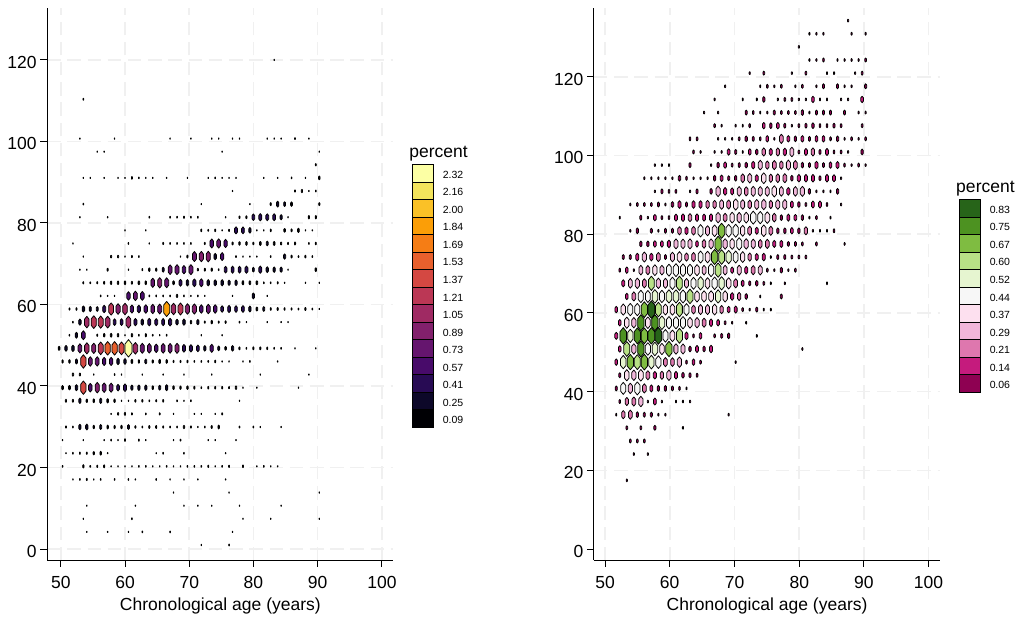

#### Supplementary Figure 5: relationship between social deficits and subjective and physiological age acceleration while additionally accounting for depression within the exposure model

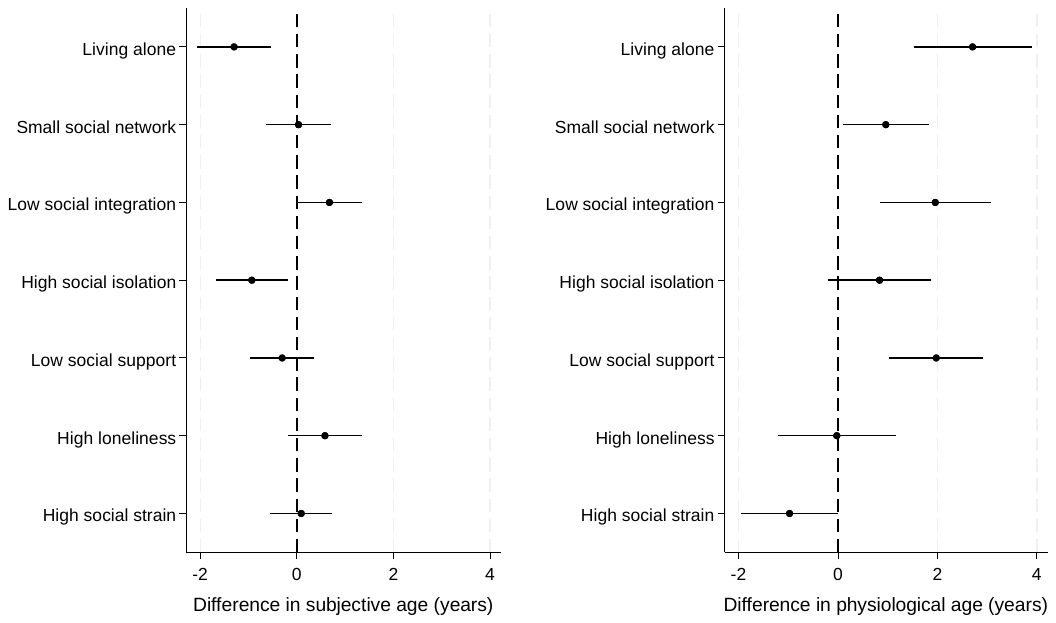

#### Supplementary Figure 6: relationship between social deficits and subjective and physiological age acceleration restricted to the same sample (n=3,878)

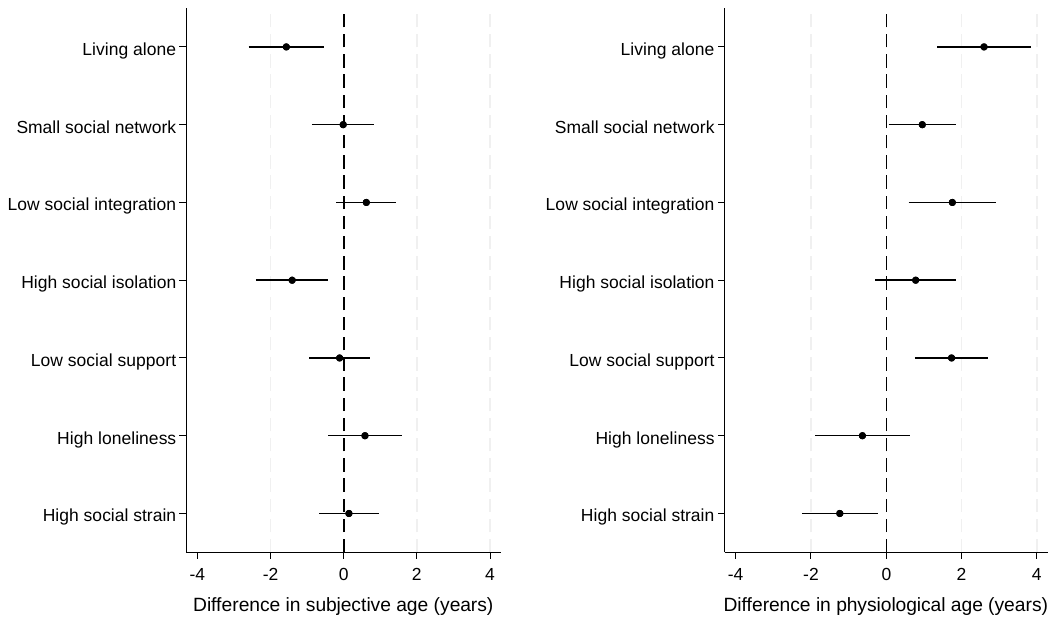

#### Supplementary Figure 7: relationship between social deficits and subjective and physiological age acceleration when removing outliers greater than 30 years above or below their chronological age (subjective age acceleration n=6,200; physiological age acceleration n=3,912)

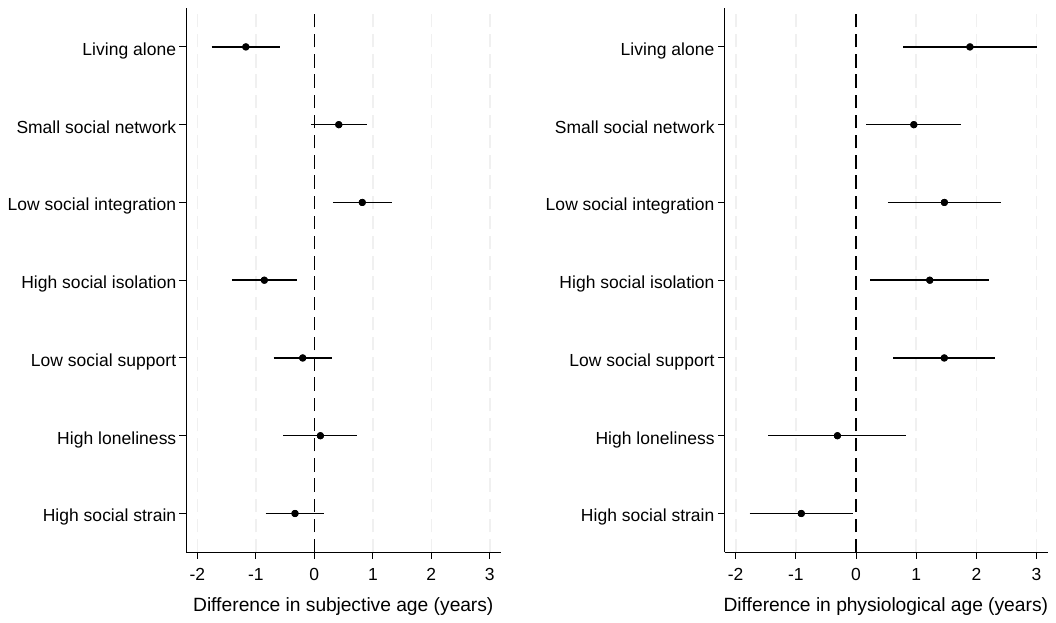

#### Supplementary Figure 8: relationship between social deficits and subjective and physiological age acceleration stratified by age (top panel, age<65; bottom panel age>=65) (For ages <65, subjective age n=3,261, physiological age n=2,118; for ages >=65, subjective age n=3,216, physiological age n=1,995)

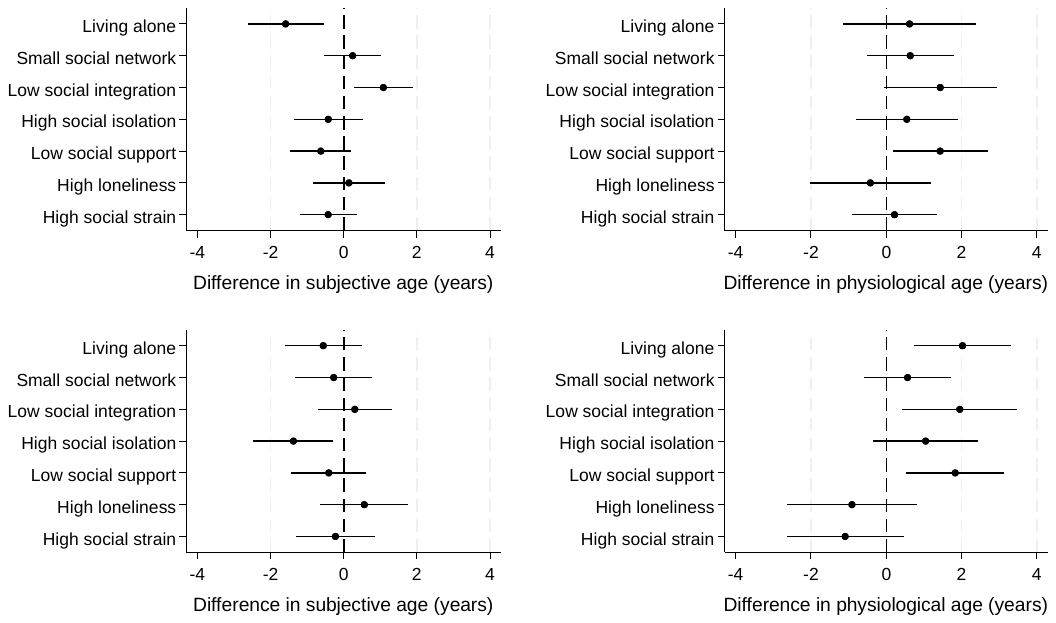

#### Supplementary Figure 9: relationship between social deficits and subjective and physiological age acceleration using outcomes from four years later (subjective age acceleration n=5,200; physiological age acceleration n=2,964)

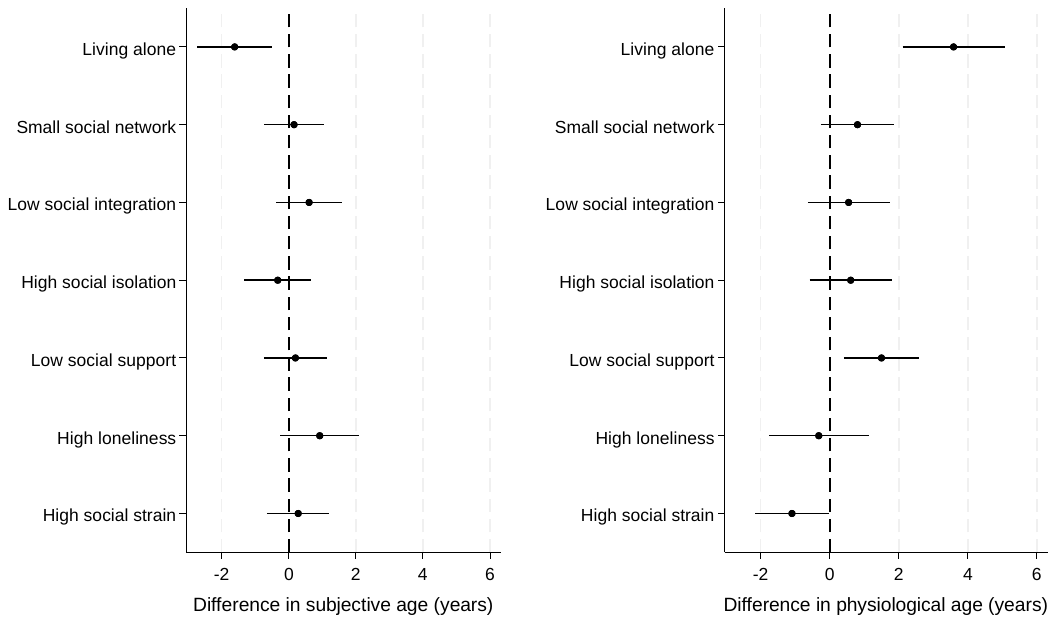

#### Supplementary Figure 10: relationship between social deficits and subjective and physiological age acceleration using an alternative computation of proportional discrepancy in age (age index-chronological age/chronological age).

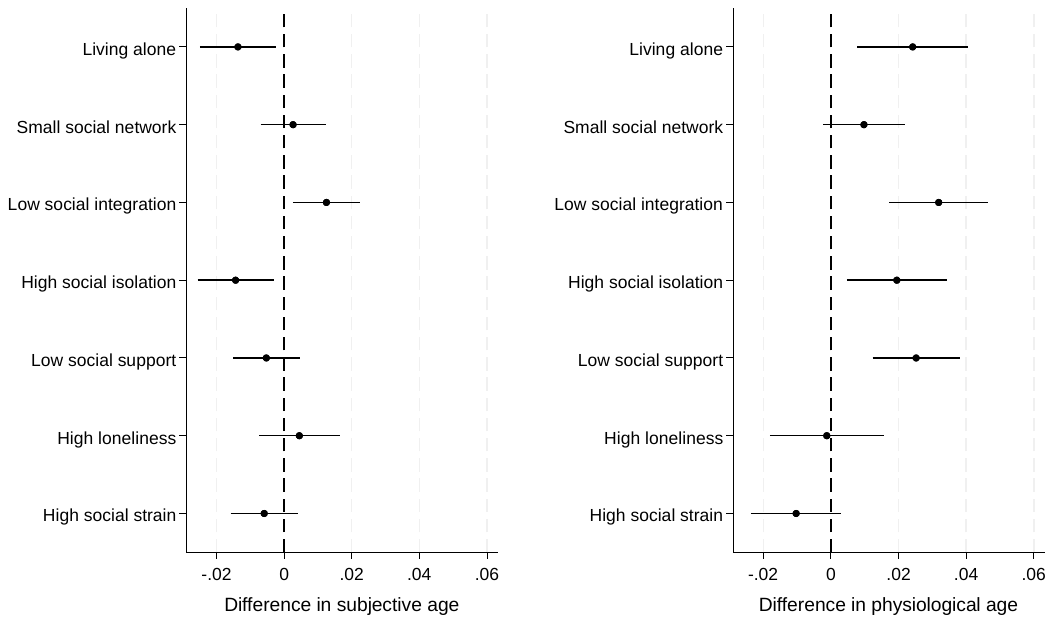

#### Supplementary Figure 11: relationship between social deficits and subjective and physiological age acceleration using continuous variables for plausibly continuous confounders

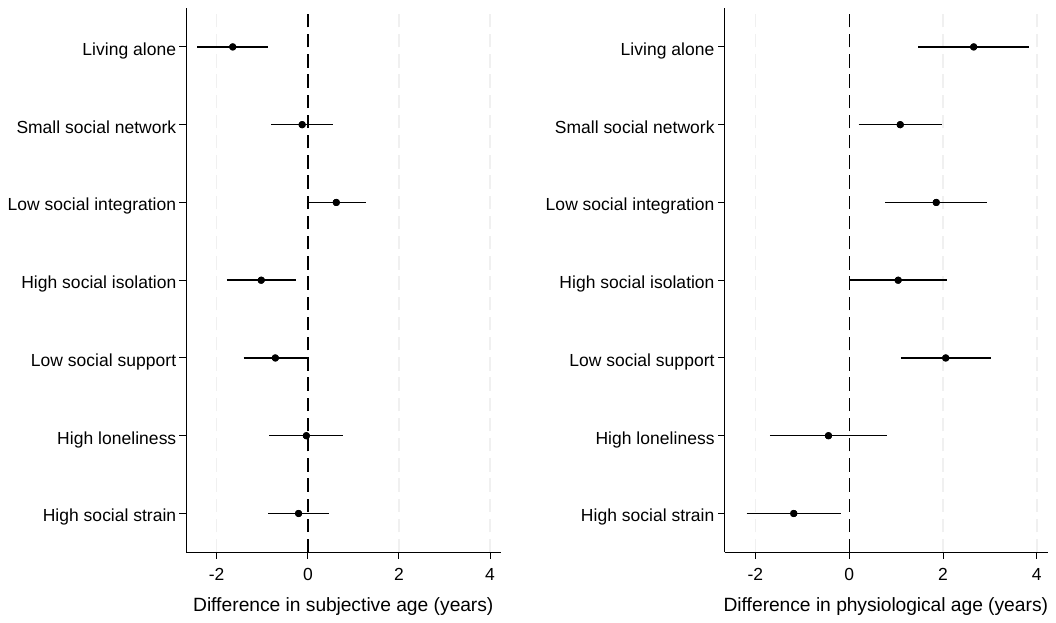
